# Diversity During Recruitment At An Internal Medicine Residency Program

**DOI:** 10.1101/2020.09.01.20186221

**Authors:** Sarwan Kumar, Deepak Gupta

## Abstract

**Background:** The right problem for graduate medical education (GME) program directors is whether diversity in their GME programs is as good as diversity in feeder entities to their GME programs. Generally, the feeder entities to GME residency programs are their affiliated medical schools. However, the specific feeder entities to GME residency programs are the unfiltered applicants’ pool who apply to these programs through Electronic Residency Application Service® (ERAS®).

**Objectives:** To analyze associations in diversity among the GME applicants, the GME interviewees and the GME residents within an internal medicine residency program assuming that unfiltered applicants’ pool is the specific feeder entity to the analyzed GME program.

**Methods:** We analyzed associations in age-group, gender, ethnicity and race diversity among the GME applicants, the GME interviewees and the GME residents within an internal medicine residency program for ERAS® 2018-2020 seasons to decipher Cramer’s V as association coefficients (“diversity scores”).

**Results:** The only significant finding was that among Not Hispanic or Latino ethnicity applications, race of ERAS® applicants had a very weak association with them being called for interviews or them becoming residents during ERAS® 2019 season as well as during the entire three-season-period (2018-2020).

**Conclusion:** Race of Not Hispanic or Latino ethnicity ERAS® applicants had a very weak association with them being called for interviews or them becoming residents at the analyzed internal medicine residency program.

## Introduction

The benefits^1^ of diversity in workforce have been made well-known for quite some time. The factors^2^ interfering with recruitment and retention of diverse workforces are discussed all the time. The unclear and abstract diversity statements^3^ have often raised eyebrows. However, the discussions have rarely focused on type III errors^4-6^ and type IV errors when quantifying diversity or factors affecting diversity in workforce. Henceforth, solving the wrong problem accidentally (type III error) or intentionally (type IV error) ignores the critical thought process needed to identify and define the right problem. For example, while testing the null hypothesis that diversity is not significantly different among physicians compared to that among general population, it is important to not only avoid (a) false positives (type I error) and (b) false negatives (type II error) but also recognize (c) type III errors among true positives of wrong null hypothesis warranting the correction of null hypothesis as a comparison of diversity among graduating medical residents with that among graduating medical students and (d) type IV errors of interpretations about true positives leading to administrative actions over-correcting or even reversing diversity among matriculating medical residents as compared to that among matriculating medical students. Therefore, it is our hypothesis (previously published as perspective paper)^7^ that the right problem for graduate medical education (GME) program directors is whether diversity in their GME programs^8^ is as good as diversity in feeder entities to their GME programs. Generally, the feeder entities to GME residency programs are their affiliated medical schools. However, the specific feeder entities to GME residency programs are the unfiltered applicants’ pool who apply to these programs through Electronic Residency Application Service® (ERAS®). More specifically, the truest feeder entities are the filtered ERAS® applicants’ pools who meet GME residency programs’ specific filtering eligibility criteria because their applications are the only ones eventually reviewed in detail by GME residency programs’ recruitment teams.

To test our hypothesis, the current study was designed to analyze associations in diversity among the GME applicants, the GME interviewees and the GME residents within an internal medicine residency program where co-author (SK) has been residency program director since 2017.

## Methods

After being approved by institutional review board as non-human participant research, ERAS® database of co-author (SK) as program director was mined and anonymously tabulated by GME applicants’, interviewees’ and residents’ age group, gender, ethnicity and race at SK’s internal medicine residency program for ERAS® 2018-2020 seasons and for this anonymized data-mining, permission from or manuscript review by ERAS® did not seem warranted. These anonymized tabulations were then analyzed to decipher Cramer’s V as association coefficients (“diversity scores”) among applicants, interviewees and residents: age-group-specific “diversity scores”, gender-specific “diversity scores”, ethnicity-specific “diversity scores” and race-specific “diversity scores” for each year as well as for the entire three-season-period (2018-2020). Although a multivariable Cox proportional hazards regression model to calculate hazard ratio would have been better, we chose Cramer’s V for its simplicity and accessibility at Vassar Stats as up to 5×5 contingency table online calculators with age group/gender/ethnicity/race categories as independent variables on its rows and applicants/interviewees/residents as “outcomes” on its columns.

Statistically, Cramer’s V tells the strength of association only when Chi-squared test evaluation of proportions is significant (P<0.05). Applicably, Cramer’s V=0 being no association among the variables was presumed to mean that age group, gender, ethnicity and race of ERAS® applicants had never been associated with them being called for interviews or them becoming residents while Cramer’s V=1 being perfect association among the variables was presumed to mean that age group, gender, ethnicity and race of ERAS® applicants had always been associated with them being called for interviews or them becoming residents. Essentially, we christened the numerical values (0-1) of only statistically significant Cramer’s V as analyzed program’s “diversity scores” with statistically significant Cramer’s V=0 meaning diversity perfectly matched among applicants, interviewees and residents while statistically significant Cramer’s V=1 meaning diversity perfectly denied among applicants, interviewees and residents.

During data-mining, it was observed that certain number of applications had missing documentations about applicants’ age, gender, ethnicity and race. Although this might have been most likely due to applicants choosing ERAS® blinded recruitment processes, we excluded all these missing documentations during our “diversity scores” analysis as tabulated in Tables 1-2(a). Additionally, as ERAS® allows free-form unstructured text along with multiple choices from among too many pre-defined options for ethnicity and race segments, we had to simplify documented ethnicity and race data before tabulating them as Tables 2 according to pre-defined options as outlined by United States Census Bureau™.^9^ While Not Hispanic or Latino ethnicity applications had their documented race, Hispanic or Latino ethnicity applications had no clearly documented race. Therefore, while ethnicity-specific “diversity scores” were possible among two ethnic groups (Table 2(a)), race-specific “diversity scores” were possible only among Not Hispanic or Latino ethnicity (Table 2(b)). Moreover, age-group >54 years (Table 1(a)) and among Not Hispanic or Latino ethnicity, American Indian and Alaskan Native alone race (Table 2(b)) were also excluded due to their almost zero numbers.

**Table 1(a):** Age-Group-Specific “Diversity Scores” During Recruitment At An Internal Medicine Residency Program In 2018-2020

| AGE GROUP | <25<br>years | 25-34<br>years | 35-44<br>years | 45-54<br>years | “Diversity<br>Score”<br>Cramer’s<br>V<br>(P Value) | EXCLUDED |  |
| --- | --- | --- | --- | --- | --- | --- | --- |
|  |  |  |  |  |  | >54<br>years | Missing<br>Data |
| 2018 |  |  |  |  |  |  |  |
| Applicants<br>(n=3294) | 254<br>(8%) | 2822<br>(86%) | 191<br>(6%) | 18<br>(1%) | 0.03<br>(0.62) | 0<br>(0%) | 9<br>(0%) |
| Interviewees<br>(n=107) | 10<br>(9%) | 94<br>(88%) | 3<br>(3%) | 0<br>(0%) |  | 0<br>(0%) | 0<br>(0%) |
| Residents<br>(n=13) | 2<br>(15%) | 11<br>(85%) | 0<br>(0%) | 0<br>(0%) |  | 0<br>(0%) | 0<br>(0%) |
| 2019 |  |  |  |  |  |  |  |
| Applicants<br>(n=3820) | 300<br>(8%) | 3262<br>(85%) | 233<br>(6%) | 23<br>(1%) | 0.03<br>(0.23) | 0<br>(0%) | 2<br>(0%) |
| Interviewees<br>(n=128) | 6<br>(5%) | 119<br>(93%) | 3<br>(2%) | 0<br>(0%) |  | 0<br>(0%) | 0<br>(0%) |
| Residents<br>(n=12) | 0<br>(0%) | 12<br>(100%) | 0<br>(0%) | 0<br>(0%) |  | 0<br>(0%) | 0<br>(0%) |
| 2020 |  |  |  |  |  |  |  |
| Applicants<br>(n=3881) | 279<br>(7%) | 3266<br>(84%) | 236<br>(6%) | 23<br>(1%) | 0.02<br>(0.89) | 2<br>(0%) | 75<br>(2%) |
| Interviewees<br>(n=138) | 12<br>(9%) | 117<br>(85%) | 7<br>(5%) | 0<br>(0%) |  | 0<br>(0%) | 2<br>(1%) |
| Residents<br>(n=12) | 1<br>(8%) | 11<br>(92%) | 0<br>(0%) | 0<br>(0%) |  | 0<br>(0%) | 0<br>(0%) |
| 2018-2020 |  |  |  |  |  |  |  |
| Applicants<br>(n=10995) | 833<br>(8%) | 9350<br>(85%) | 660<br>(6%) | 64<br>(1%) | 0.02<br>(0.17) | 2<br>(0%) | 86<br>(1%) |
| Interviewees<br>(n=373) | 28<br>(8%) | 330<br>(88%) | 13<br>(3%) | 0<br>(0%) |  | 0<br>(0%) | 2<br>(1%) |
| Residents<br>(n=37) | 3<br>(8%) | 34<br>(92%) | 0<br>(0%) | 0<br>(0%) |  | 0<br>(0%) | 0<br>(0%) |

**Table 1(b):**
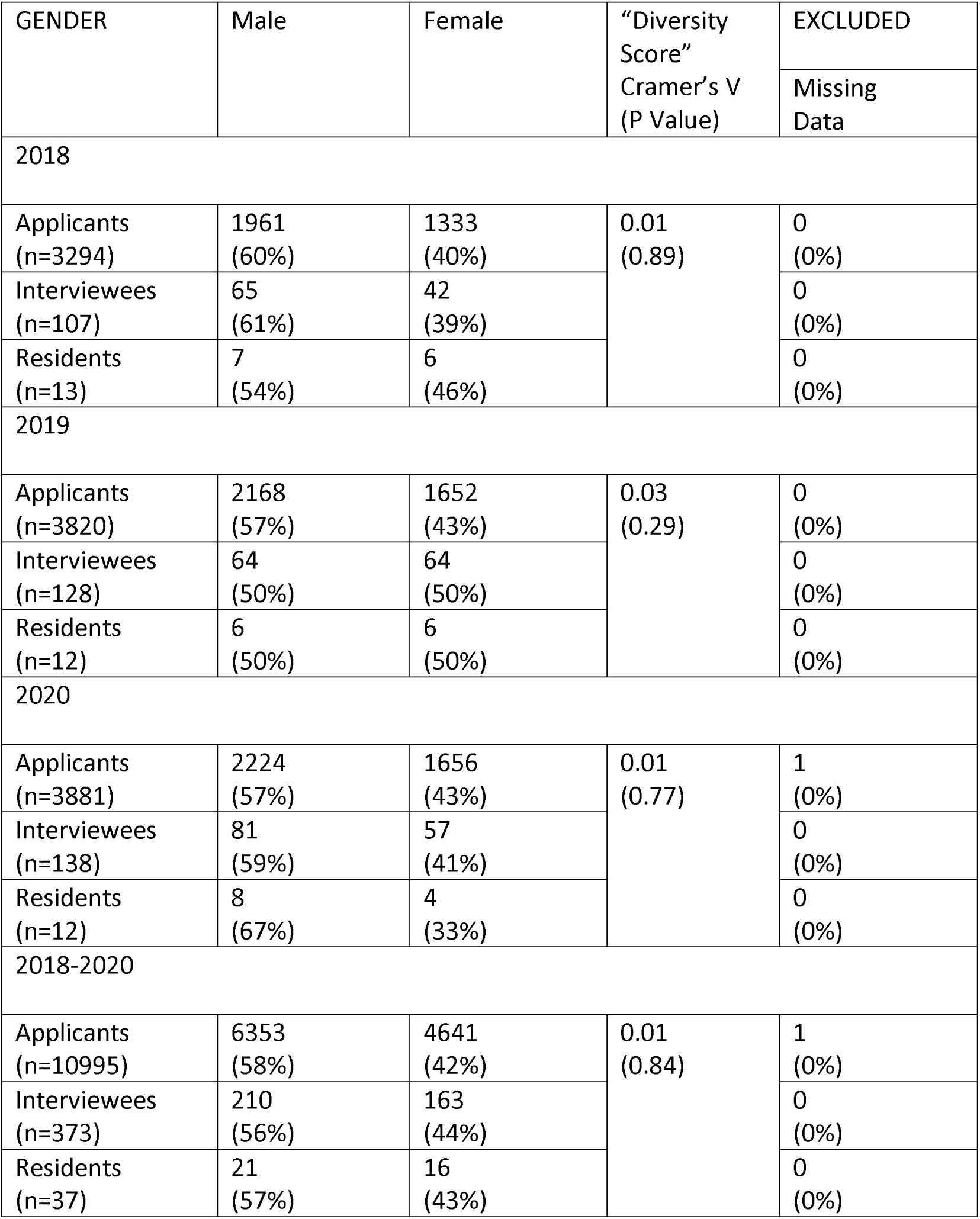
Gender-Specific “Diversity Scores” During Recruitment At An Internal Medicine Residency Program In 2018-2020

**Table 2(a):**
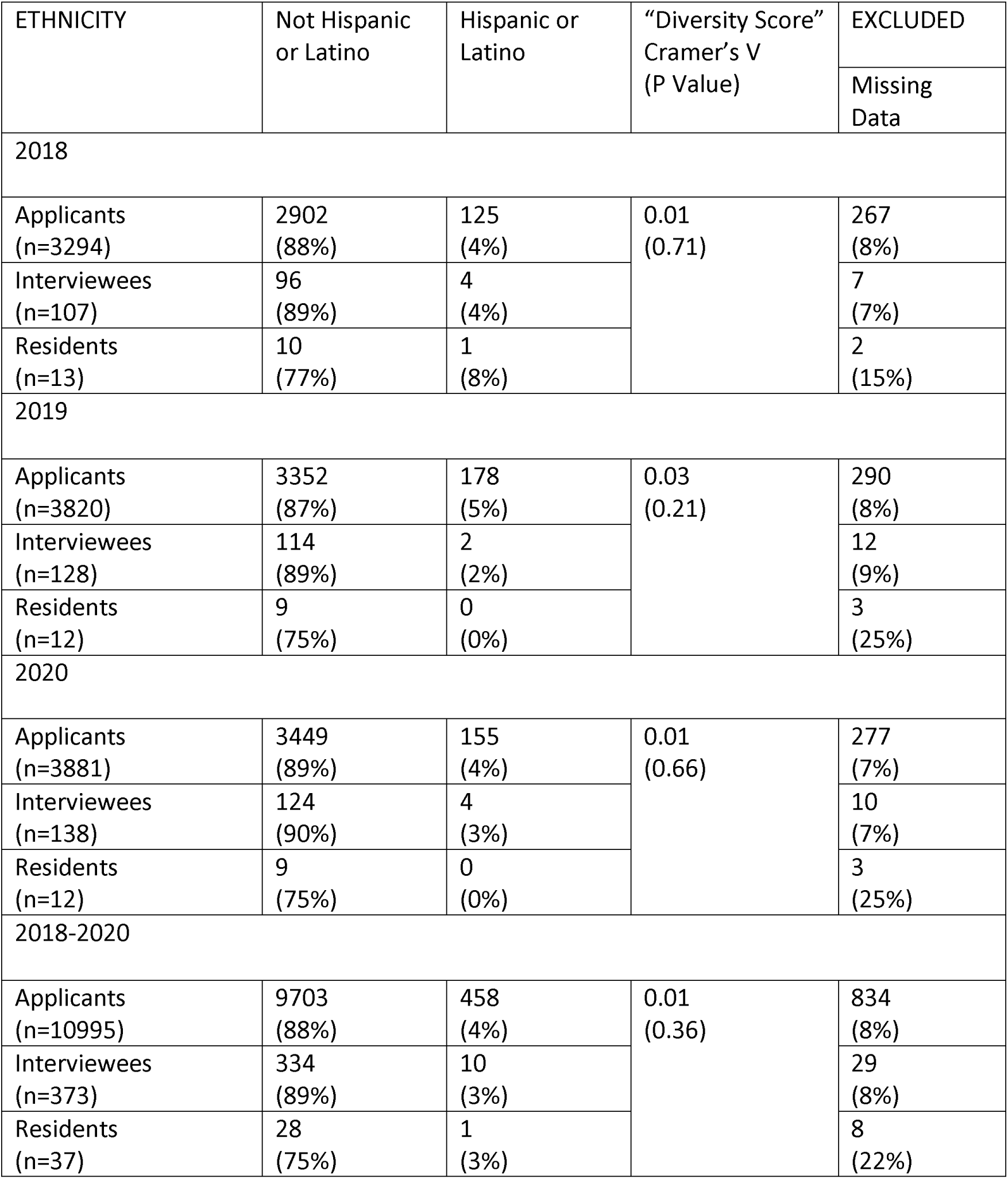
Ethnicity-Specific “Diversity Scores” During Recruitment At An Internal Medicine Residency Program In 2018-2020

| ETHNICITY | Not Hispanic<br>or Latino | Hispanic or<br>Latino | “Diversity Score”<br>Cramer’s V<br>(P Value) | EXCLUDED |
| --- | --- | --- | --- | --- |
|  |  |  |  | Missing<br>Data |
| 2018 |  |  |  |  |
| Applicants<br>(n=3294) | 2902<br>(88%) | 125<br>(4%) | 0.01<br>(0.71) | 267<br>(8%) |
| Interviewees<br>(n=107) | 96<br>(89%) | 4<br>(4%) |  | 7<br>(7%) |
| Residents<br>(n=13) | 10<br>(77%) | 1<br>(8%) |  | 2<br>(15%) |
| 2019 |  |  |  |  |
| Applicants<br>(n=3820) | 3352<br>(87%) | 178<br>(5%) | 0.03<br>(0.21) | 290<br>(8%) |
| Interviewees<br>(n=128) | 114<br>(89%) | 2<br>(2%) |  | 12<br>(9%) |
| Residents<br>(n=12) | 9<br>(75%) | 0<br>(0%) |  | 3<br>(25%) |
| 2020 |  |  |  |  |
| Applicants<br>(n=3881) | 3449<br>(89%) | 155<br>(4%) | 0.01<br>(0.66) | 277<br>(7%) |
| Interviewees<br>(n=138) | 124<br>(90%) | 4<br>(3%) |  | 10<br>(7%) |
| Residents<br>(n=12) | 9<br>(75%) | 0<br>(0%) |  | 3<br>(25%) |
| 2018-2020 |  |  |  |  |
| Applicants<br>(n=10995) | 9703<br>(88%) | 458<br>(4%) | 0.01<br>(0.36) | 834<br>(8%) |
| Interviewees<br>(n=373) | 334<br>(89%) | 10<br>(3%) |  | 29<br>(8%) |
| Residents<br>(n=37) | 28<br>(75%) | 1<br>(3%) |  | 8<br>(22%) |

**Table 2(b):**
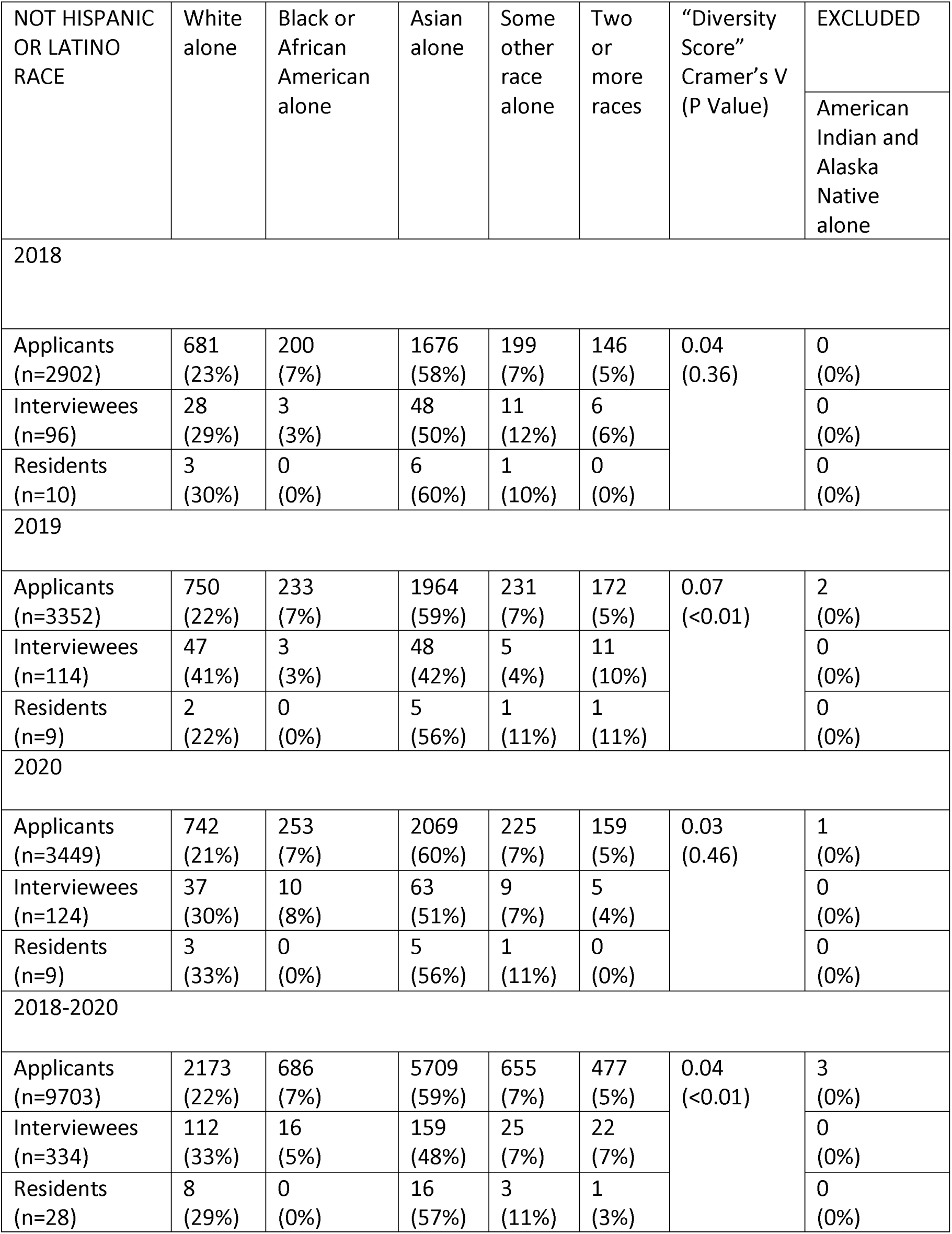
Not-Hispanic-Or-Latino-Race-Specific “Diversity Scores” During Recruitment At An Internal Medicine Residency Program In 2018-2020

| NOT HISPANIC<br>OR LATINO<br>RACE | White<br>alone | Black or<br>African<br>American<br>alone | Asian<br>alone | Some<br>other<br>race<br>alone | Two<br>or<br>more<br>races | “Diversity<br>Score”<br>Cramer’s V<br>(P Value) |  |
| --- | --- | --- | --- | --- | --- | --- | --- |
|  |  |  |  |  |  |  | American<br>Indian and<br>Alaska<br>Native<br>alone |
| 2018 |  |  |  |  |  |  |  |
| Applicants<br>(n=2902) | 681<br>(23%) | 200<br>(7%) | 1676<br>(58%) | 199<br>(7%) | 146<br>(5%) | 0.04<br>(0.36) | 0<br>(0%) |
| Interviewees<br>(n=96) | 28<br>(29%) | 3<br>(3%) | 48<br>(50%) | 11<br>(12%) | 6<br>(6%) |  | 0<br>(0%) |
| Residents<br>(n=10) | 3<br>(30%) | 0<br>(0%) | 6<br>(60%) | 1<br>(10%) | 0<br>(0%) |  | 0<br>(0%) |
| 2019 |  |  |  |  |  |  |  |
| Applicants<br>(n=3352) | 750<br>(22%) | 233<br>(7%) | 1964<br>(59%) | 231<br>(7%) | 172<br>(5%) | 0.07<br>($<0.01$) | 2<br>(0%) |
| Interviewees<br>(n=114) | 47<br>(41%) | 3<br>(3%) | 48<br>(42%) | 5<br>(4%) | 11<br>(10%) |  | 0<br>(0%) |
| Residents<br>(n=9) | 2<br>(22%) | 0<br>(0%) | 5<br>(56%) | 1<br>(11%) | 1<br>(11%) |  | 0<br>(0%) |
| 2020 |  |  |  |  |  |  |  |
| Applicants<br>(n=3449) | 742<br>(21%) | 253<br>(7%) | 2069<br>(60%) | 225<br>(7%) | 159<br>(5%) | 0.03<br>(0.46) | 1<br>(0%) |
| Interviewees<br>(n=124) | 37<br>(30%) | 10<br>(8%) | 63<br>(51%) | 9<br>(7%) | 5<br>(4%) |  | 0<br>(0%) |
| Residents<br>(n=9) | 3<br>(33%) | 0<br>(0%) | 5<br>(56%) | 1<br>(11%) | 0<br>(0%) |  | 0<br>(0%) |
| 2018-2020 |  |  |  |  |  |  |  |
| Applicants<br>(n=9703) | 2173<br>(22%) | 686<br>(7%) | 5709<br>(59%) | 655<br>(7%) | 477<br>(5%) | 0.04<br>($<0.01$) | 3<br>(0%) |
| Interviewees<br>(n=334) | 112<br>(33%) | 16<br>(5%) | 159<br>(48%) | 25<br>(7%) | 22<br>(7%) |  | 0<br>(0%) |
| Residents<br>(n=28) | 8<br>(29%) | 0<br>(0%) | 16<br>(57%) | 3<br>(11%) | 1<br>(3%) |  | 0<br>(0%) |

## Results

As shown in Tables 1-2(a), age-group-specific, gender-specific and ethnicity-specific “diversity scores” were not statistically significant and thus their Cramer’s V (level of association’s strength) were irrelevant during ERAS® 2018-2020 seasons at the analyzed internal medicine residency program. Among race-specific “diversity scores” among Not Hispanic or Latino ethnicity applications as shown in Table 2(b), P-value was significant (<0.01) during ERAS® 2019 season as well as during the entire three-season-period (2018-2020) but Cramer’s V (“diversity scores”) in both instances were very close to 0 as shown in Table 2(b) (0.07 for 2019 season; 0.04 for 2018-2020 period) indicating that race of ERAS® applicants had a very weak association with them being called for interviews or them becoming residents during ERAS® 2019 season as well as during the entire three-season-period (2018-2020).

## Discussion

The only key finding of our non-human participant research was that among Not Hispanic or Latino ethnicity applications, race of ERAS® applicants had a very weak association with them being called for interviews or them becoming residents. To validate or refute this finding in the future, we strongly urge ERAS® that instead of allowing applicants to leave age, gender, ethnicity and race segments empty, it should mandate such documentations by all applicants wherein they can always choose the pre-defined option “Decline to answer” when documenting their age, gender, ethnicity and race. Similarly, instead of free-form unstructured text, ERAS® ethnicity and race segment should have structured text format allowing only one option to choose from among pre-defined options as outlined by United States Census Bureau™.^9^ This will allow ERAS® ethnicity and race data to be more appropriately followed for tracking and aligning future “diversity scores” by individual GME programs locally and by ERAS® regionally-nationally.

During our current analysis, we could not investigate associations among the filtered ERAS® applicants’ pools during 2018-2020 seasons who had met the analyzed internal medicine residency program’s specific filtering eligibility criteria. Therefore, our current analysis had to use the unfiltered applicants’ pools who had applied to the analyzed internal medicine residency program during ERAS® 2018-2020 seasons. In the future ERAS® seasons, we are hoping to utilize our “diversity scores’ methodology by prospectively investigating association among all filtered applications (all reviewed applicants) in terms of age-group, gender, ethnicity and race with those among all scheduled interviewees. Thereafter, the analyzed internal medicine program can average the lowest rank numbers at which the ranked interviewees had matched as residents over the prior three years. Assuming that the analyzed internal medicine program matriculates 12 residents annually and the three-year averaged number for the lowest rank at which the ranked interviewees had matched is 50, the “diversity scores” methodology can be further applied to investigate association among all scheduled interviewees with the first 50-ranked interviewees regarding their age-group, gender, ethnicity and race. Essentially, compared to current “diversity scores” methodology retrospectively investigating association among unfiltered applicants with scheduled interviewees with matched residents, the future age-group-specific, gender-specific, ethnicity-specific and race-specific “diversity scores” may attempt to prospectively correct inadvertent association among age-group, gender, ethnicity and race of filtered-and-reviewed applicants with scheduled interviewees with first 50-ranked interviewees because these are the ones which are under primary control of residency programs’ recruitment teams. Alternatively, diversity can be simply realigned prospectively with our diversity realignment score as explained in our supplementary excel file of simulated data along with its explanation in our supplementary video file whose detailed and expanded version can be additionally viewed on YouTube.^10^

There were few additional limitations to our retrospective analysis. A multivariable Cox proportional hazards regression model to calculate hazard ratio as applicants evolve to being interviewees to finally become residents may have been better than Cramer’s V for determining the effect of each independent variable while controlling the effect of other independent variables (age-group/gender/ethnicity/race). Data imputation approach may have prevented exclusions of missing documentations about ethnicity/race. Our single-institution analysis for brief period of three years needs further validation by multi-institutional prospective studies utilizing the above-mentioned better statistical methods. Once our results have been validated by multi-institutional prospective studies, our “diversity scores” methodology may turn out to be a simple method to guide GME program directors during their attempts to ensure age-group-specific, gender-specific, ethnicity-specific and race-specific diversity among the recruited GME workforce in the future.

## Conclusion

Race of Not Hispanic or Latino ethnicity ERAS® applicants had a very weak association with them being called for interviews or them becoming residents at the analyzed internal medicine residency program.

## Supporting information

Diversity Realignment Score Example (Simulation)

Diversity Realignment Score Video (Simulation)

Non-Human Participant Research

## Data Availability

All data as tabulated in the manuscript is available for review.

## Acknowledgements

The authors are thankful to Dr Ronald Thomas, Children’s Research Center of Michigan, Wayne State University, for overseeing the statistical analysis for this project.

