## Supplementary material for "Diversity During Recruitment At An Internal Medicine Residency Program": Non-Human Participant Research

### WAYNE STATE UNIVERSITY

**INSTITUTIONAL REVIEW BOARD**
**87 E. Canfield, 2nd Floor**
**Detroit, Michigan 48201**
**www.irb.wayne.edu**

#### NOTICE OF IRB ADMINISTRATIVE DETERMINATION: NON-HUMAN PARTICIPANT RESEARCH

**TO:** Sarwan Kumar, MD

**FROM:** Heather Park-May

**DATE:** March 5, 2020

**RE:** Correlating Diversity Among Matriculated Residents with Diversity Among Interviewed Applicants and Diversity among GME Applicants

**WSU IRB Number** 2020 036

---

Materials concerning the above-referenced proposal was initially received by the Wayne State University Institutional Review Board Administration Office via Human Participant Research Determination Tool on March 4, 2020.

A determination has been made that this project does not constitute human participant research according to the definition codified in the Common Rule at 45 CFR 46 and FDA regulations. This means that IRB review and oversight is not required for this project. We recommend you keep this memo for your records.

The project also does not involve individuals who would receive a test article (drug or device) as participants and therefore the FDA regulations do not apply. Thus, this project does not require review or approval by the Wayne State University Institutional Review Board.

**Please be aware that projects involving identifiable Personal Health Information (PHI) which the IRB has determined does not meet the definition of research must still adhere to HIPAA regulations, and may require authorization from the covered entity's privacy officer.**

Please note that changes to the study plan may impact this determination as to whether the project constitutes Human Participant Research. Please contact the IRB Administration office if there are changes to the study plan that may affect this determination.

H. Park-May
Wayne State University, IRB Administration Office

03/05/2020
Date

**WAYNE STATE UNIVERSITY**

**IRB Administration Office**
87 E. Canfield, Second Floor
Office (313) 577-1628
Detroit, MI 48201
http://irb.wayne.edu/index.php

### Human Participant Research Determination Tool

The regulatory requirement for IRB review under the Common Rule applies to research that is "a systematic investigation, including research development, testing and evaluation, designed to develop or contribute to generalizable knowledge." Only research meeting the definition of Human Participant Research (HPR) or research for which the FDA regulations apply requires IRB review and IRB oversight.

This tool should be used for determining when a project requires IRB review and approval. Use this tool to determine if a project meets the regulatory definition of research requiring IRB review in number 1 and 2.

**Note**: Human participation research is defined by two Federal regulatory agencies (DHHS and FDA). This tool will help to determine whether your activity is subject to one or both Federal agency regulations. If the activity is determined to be human participant research according to DHHS regulation as defined in sections A and B, that does not necessarily mean it is also subject to FDA regulation, and vice versa. Separate determinations should be made.

If assistance is needed,or if written documentation from the IRB office is required, complete the **entire** form and submit the form and any relevant supporting documents (e.g., grant, protocol, data collection tools) to the IRB administration office, or email it to . Please do not submit handwritten documents to the IRB office.

**HPR Determination Number** 2020 036
IRB Use ONLY

#### Project Information:

Complete this section if you will be requesting assistance from the IRB Administration Office in making your Human Participation Research determination. Otherwise continue on to number 1 to begin the determination tool.

| | | | | |
|---|---|---|---|---|
| Project Title: | **Correlating Diversity Among Matriculated Residents With Diversity Among Interviewed Applicants And Diversity Among GME Applicants** | | | |
| Name of person conducting the project: | **Sarwan Kumar** | Title: | **MD** | Date: 03/04/2020 |
| Status: **Select all that apply** | ☒ Wayne State Faculty ☐ WSU Graduate Student ☐ WSU Undergraduate Student<br>☐ DMC Staff ☐ Karmanos Staff ☐ J. D. Dingell VAMC Staff<br>☐ Resident/Fellow/Trainee ☐ Other: _____ | | | |
| Division or College: | Crittenton Hospital | Campus Address: | Program Director<br>Internal Medicine Residency Program<br>Wayne State University/Ascension Providence Rochester Hospital<br>1101 W. University Drive, 2 South<br>Rochester, MI 48307 | |
| Department: | Internal Medicine | Email Address: | **** | |
| Alternate or Home Address: | ☒ N/A | Phone: | Office (248) 601-4805<br>Fax (248) 601-4908 | |

| Faculty Sponsor/ Supervisor for this Project: | Name:<br><br>Email:<br>☒ I do not have a Faculty Sponsor/Supervisor | Phone: | ( ) |
| --- | --- | --- | --- |
| | | Title: | |
| Form completed by: | Deepak Gupta MD | E-mail: | |

| Check ALL that apply: | ☒ Behavioral, social, education, non-medical research<br>☒ Medical research |
| --- | --- |

**Provide a description of the project with enough detail for the determination. Enter "N/A" where appropriate.**

Describe the purpose, study question, study objectives or aims for this project:

**The study question is whether diversity (age-gender-race-ethnicity) among matriculated residents in Internal Medicine Residency Program at Wayne State University/Ascension Providence Rochester Hospital over three years (2018-2020) correlates with diversity (age-gender-race-ethnicity) among interviewed applicants and GME applicants for Internal Medicine Residency Program at Wayne State University/Ascension Providence Rochester Hospital for the same three years (via ERAS 2018-2020). The ERAS portal based database will be accessed for all the GME applicants as well as all the interviewed applicants for three years (2018-2020) so as to non-identifiably tabulate their age group, gender, race and ethnicity and thereafter the matriculated residents for the same three years (2018-2020) will also be non-identifiably tabulated per their age group, gender, race and ethnicity. Thereafter, the GME applicants' characteristics' proportions regarding their age group, gender, race and ethnicity will be correlated to the interviewed applicants' characteristics' proportions regarding their age group, gender, race and ethnicity which in turn will be correlated to the matriculated residents' characteristics' proportions regarding their age group, gender, race and ethnicity. These Pearson correlation coefficients (-1 to +1) for age group, gender, race and ethnicity will be able to inform how diversity among matriculated residents is correlating with diversity among interviewed applicants and thereto with diversity among GME applicants in Internal Medicine Residency Program at Wayne State University/Ascension Providence Rochester Hospital.**

Describe how the results will be used including any plans for presentation or publication:

**The results of this program evaluation/quality improvement/quality assurance project will be presented at local/regional/national conferences and/or published at peer-reviewed journals so as to share this local data's results with medical educators and clinical researchers globally.**

State the location(s) where research activities will take place:

**Internal Medicine Residency Program at Wayne State University/Ascension Providence Rochester Hospital**

Describe the participants (if applicable) for the project:

The GME applicants' (along with interviewed applicants' and matriculated residents') characteristics accessible directly from ERAS portal based database for Internal Medicine Residency Program at Wayne State University/Ascension Providence Rochester Hospital

Describe the data/information that would be collected for the study and the source(s) of that data:

The GME applicants' (along with interviewed applicants' and matriculated residents') characteristics will be non-identifiably tabulated only for their age group, gender, race and ethnicity.

Describe how data will be obtained (e.g. survey, interview, observation, testing, review of existing records, etc.):

The GME applicants' (along with interviewed applicants' and matriculated residents') characteristics will be accessed directly from ERAS portal based database for Internal Medicine Residency Program at Wayne State University/Ascension Providence Rochester Hospital

Describe whether or not the data will include individually identifying information (e.g. name, DOB, MRN, email address, other codes; etc.):

Non-identifiable tabulations of age group, gender, race, ethnicity of GME applicants (along with interviewed applicants and matriculated residents) for three years 2018-2020

Could the identities of participants be known to, or be readily ascertained by the investigators? ☐ Yes ☒ No

**Instructions for submitting to WSU IRB for an official IRB determination:**

In addition to providing a complete description above and completing all sections of the determination tool below, please submit any relevant supporting documents (e.g., grant, proposal, data collection tools etc.) with this tool to the IRB administration office, or as an email to the IRB Education Coordinator for assistance in making the determination.

IRB Administration Office Staff Contact Information: http://irb.wayne.edu/ContactUs.php

#### Determination Tool:

1. **Does the activity involve:**
   a. Prospective collection of information or bio-specimens through intervention or interaction and uses, studies, or analyzes the information or bio-specimens? ☐ Yes ☒ No
   b. The collection or use of individually identifiable and private information? ☐ Yes ☒ No
      -If you answered yes to either 1a or 1b then your activity involves human participants.

2. **Does the activity involve:**
   a. As systematic investigation ☐ Yes ☒ No
   b. Is the activity designed to develop or contribute to generalizable knowledge? ☐ Yes ☒ No
      -If you answered yes to either 2a or 2b then your activity involves research.

**Your activity requires IRB review if it involves both human participants AND research as determined above. Please see definitions below to help interpret some of the terms mentioned in this tool.**

**Does the activity require IRB review under the FDA Regulations?**

3. **If any of the following apply, your project will require IRB review under FDA regulations. Check all that apply.**
   - ☐ In the United States: The use of a drug in one or more persons other than the use of an approved drug in the course of medical practice
   - ☐ In the United States: The use of a device in one or more persons or human specimens that evaluates the safety or effectiveness of that device
   - ☐ Data regarding participants or control participants submitted to or held for inspection by FDA
   - ☐ Data regarding the use of a device on human specimens (identified or unidentified) submitted to or held for inspection by the FDA

#### Definitions

**Intervention:** Physical procedure by which information or bio-specimens are gathered and manipulations of the participant or the participant's environment that are performed for research purposes (45 CFR 46.102).

**Interaction:** Communication or interpersonal contact between an investigator and the participant (including electronic interaction) (according to OHRP).

**Individually Identifiable:** The identity of the participant is or may readily be ascertained by the investigator or those associated with the information (according to OHRP).

**Private Information:** Information provided for specific purposes by an individual if the individual can reasonably expect that no observation or recording is taking place, and information provided will not be made public (e.g., medical or psychological information (According to OHRP).

**Systematic Investigation:** Activity that involves development, testing, evaluation, and data collection with either quantitative or qualitative data analysis to search for information and/or to answer a question (WSU 16-1 Glossary)

**Generalizable Knowledge:** Activity that draws general conclusions (knowledge gained may apply to other populations outside of the study), informs policy, or is universally or widely applicable.

- **Your project requires IRB review if #1 determined that your study involves human participants, AND #2 determined your study involves research.**
- **Your project requires IRB review under the FDA regulations if you check any of the conditions in #3**

**Useful Tips:**

1. If your research involves the use of de-identified or coded bio-specimens, include a letter of support from the department providing the bio-specimens that confirms that the bio-specimens provided will be stripped of all identifiable information before you receive it.
2. If your research involves the use of de-identified data, make it clear in your project description on page 2 that all identifiable information will be removed before you receive the data.

**Next Steps:**

- ✓ If by the use of this tool, you have determined that the project does not require IRB review, you do not need to submit this form to the IRB office. Add the project title and name of the person completing the project, their title and the date the tool was completed to the first page, and retain this tool in your files to document this determination.
- ✓ If you have determined that IRB review is required, IRB approval must be obtained **before** conducting any human participant research activities. Visit the WSU IRB website for additional information and the forms required for a new submission: http://irb.wayne.edu/. with any questions that come up along the way.
- ✓ The Self-Assessment/Pre-Review tool is useful for completing before submitting to the IRB. The tool walks you through the submission requirements based on the many different scenarios unique to each study. This tool can be used as part of the IRB pre-review or used solely by the submitter. The tool helps submitters know what documents and forms are required for IRB, as well as the type of IRB reviews and a Check for Completeness Assessment. The self-pre-review is not a required IRB form.
- ✓ If you are unsure as to whether or not this project is human participant research requiring IRB review, then complete the Project Information section on the first page and submit this form and any relevant supporting documents to the IRB office.
- ✓ If there are any modifications to your project that could change this determination, please complete this tool again. Submit the appropriate application to the IRB if changes to your project determine that human participant research is involved according to this tool which is based on the federal regulations.

---

#### WSU IRB Determination:

(To be completed by IRB Administration)

☑ **Not Human Participant Research - IRB review is not required**

- ☐ Case Report

  **Note:** IRB approval is required if the case report involves more than three cases.
- ☐ Course Related Activities

  **Note:** IRB Approval is required if a student is involved in an activity designed to teach research methodologies and the instructor or student wishes to conduct further investigation and analyses in order to contribute to scholarly knowledge.
- ☐ Decedents: Research limited to death records, autopsy materials or cadaver specimens.

**Note:** IRB approval is required if decedent information contains psychotherapy notes, or information related to HIV, mental health, genetic testing or drug or alcohol abuse

☐ Journalism/Documentary Activities

**Note:** IRB approval may be required when journalists conduct activities normally considered scientific research intended to develop generalizable knowledge (e.g. systematic research, surveys, and/or interviews that are intended to test theories or develop models).

☐ Oral History

**Note:** IRB approval is required when the activities are intended to develop generalizable conclusions (e.g., that serve as data collection intended to test economic, sociological, or anthropological models/theories)

☒ Program Evaluation/Quality Improvement/Quality Assurance Activities

**Note:** Investigators conducting QI/QA projects should ensure that they have received approval from any applicable committees within their department or the site in which the activity will occur

☐ Public Use Datasets

**Note:** IRB approval is required for the use of restricted use data, if a proposal is required to obtain the dataset, or if a data use agreement is involved.

☐ Coded or De-Identified Private Information and/or Human Biological Specimens

**Note:** IRB approval is required if the information being collected could enable the investigator to identify or readily ascertain the identity of the individual whom the private information or specimens belongs to.

☐ Other Project type not considered to be HPR

Describe:

☐ **Exempt IRB review is required**

Rationale:

☐ **Expedited IRB review is required**

Rationale:

☐ **Full Board IRB review is required**

Rationale:

Comments: [illegible]

Authorized IRB Reviewer Signature: [illegible] Date 3/5/20

Printed name: [illegible]

DATA COLLECTION SHEET

| S. No. | ERAS YEAR<br>ERAS 2018/<br>ERAS 2019/<br>ERAS 2020 | AGE GROUP<br><25 years/<br>25-34 years/<br>35-44 years/<br>45-54 years/<br>55-64 years/<br>>65 years | GENDER<br>Male/<br>Female/<br>Unknown | Race | Ethnicity | STATUS<br>Applicant/<br>Interviewee/<br>Matriculant |
|---|---|---|---|---|---|---|
